# Estimating the super-spreading rate at workplaces using bluetooth technology

**DOI:** 10.1101/2021.03.04.21252550

**Authors:** Alexei Vazquez, Maximilian Staebler, Alexander Khanin, Daniel Lichte, Eva Brucherseifer

**Affiliations:** German Aerospace Center (DLR), Institute for the Protection of Terrestrial Infrastructures, Rathausallee 12, 53757 Sankt Augustin, Germany

**Author notes:** Equal contribution (Alexei Vazquez, Maximilian Staebler).

## Abstract

Workplaces deploy internal guidelines to remain operational during the ongoing COVID-19 pandemic. It is challenging to assess whether those interventions will prevent super-spreading events, where an infected individual transmits the disease to 10 or more secondary cases. Here we provide a model of infectious disease at the level of a workplace to address that problem. We take as input proximity contact records based on bluetooth technology and the infectious disease parameters from the literature. Using proximity contact data for a case-study workplace and an infection transmission model, we estimate the SARS-CoV-2 transmission rate as 0.014 per proximity contact, going up to 0.041 for the SARS-CoV-2 B.1.1.7 variant first detected in the UK. Defining super-spreading as events with 10 or more secondary infections, we obtain a super-spreading event rate of 2.3 per 1000 imported SARS-CoV-2 cases, rising up to 13.7 for SARS-CoV-2 B.1.1.7. This methodology provides the means for workplaces to determine their internal super-spreading rate or other infection related risks.

## Introduction

The COVID-19 pandemic has tested our ability to control an infectious disease outbreak at multiple scales ^1^. Air traffic was almost completely shut down at the earlier stages ^2^. Lockdown or curfews followed in cities, regions and countries where the COVID-19 incidence rose to alarming levels ^3^. After the airborne nature of the SARS-CoV-2 (SARS2) virus was confirmed, mask use and social distancing were introduced ^4^. These non-pharmaceutical interventions were successful, where and when applied, reducing the reproductive number and mortality ^5^. Vaccinations have started in several countries but we still have a challenging year ahead. The relaxation of lockdowns has lead to secondary outbreaks worldwide. More alarmingly, a new variant of SARS2 has been identified in the United Kingdom (lineage B.1.1.7 ^6^) with a higher reproductive number ^7, 8^. The SARS2-B.1.1.7 variant is already spreading worldwide.

The economic and psychological impacts of lockdowns have changed our response to the pandemic ^9^. The alternative to lockdowns is a distributed approach where workplaces deploy containment measures to remain operational. In the distributed scenario workplaces, households and individuals are responsible to carry out the containment measures. Each workplace must do their part for this distributed approach to work. To achieve this goal, workplaces need support with knowledge and tools. The focus of most studies on COVID-19 transmission dynamics has been either at the city/country wide level ^10–16^ or at the level of aerosol transmission in one room ^17, 18^.More work needs to be done at the level of workplaces, taking into account the temporal network of proximity contacts.

Here we investigate the generation of SARS2 outbreaks within a workplace, using as input proximity contact data gathered with bluetooth technology. Our main results are an estimate of the SARS2 rate of transmission per proximity contact, a generative model to simulate infectious disease outbreaks within workplaces, estimates of the rate of super-spreading events per imported case and an evaluation of mask use as an example of non-pharmaceutical interventions within the workplace.

## Results

### Proximity contact alarm system

We have collected anonymous proximity contact data from a workplace. The workers wore button devices that reported a distance alert when the distance between two coworkers was less than 1.5m for 15 seconds (Fig. 1A). The buttons communicated with gateway beacons using bluetooth technology, which sent data to a backend server. Overall we recorded 21182 proximity contacts between 605 workers for a period of 44 days. The total number of contacts between any two pairs of individuals fluctuates over time, from values close to zero during weekends to as high as 20 contacts per individual during working days (Fig. 1B).

**Figure 1.**
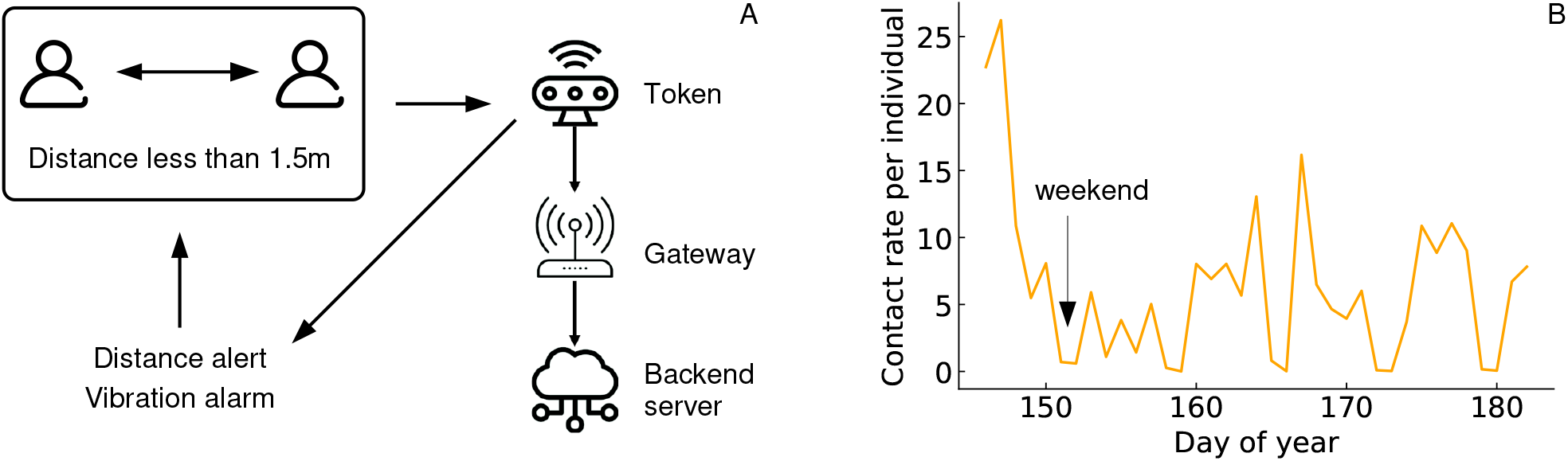
Proximity contact records. A) Schematic representation of the system recording proximity contacts. B) Time series of the contact rate per individual based on data for the case-study workplace.

The fluctuations between weekdays introduce uncertainty in the expectation of spreading outbreaks size. An individual becoming infectious on Monday has more potential to infect other individuals at the workplace than one becoming infectious on Friday and going home during the infectious period. Furthermore, the correlation of events happening within the same day will be shown to be important below.

### Disease transmission model

The proximity contact records provide a temporal network to model the spread of airborne viruses. Using the language of infectious diseases, a primary case is an infected individual who is the focus of our attention and secondary cases are individuals becoming infected after contact with the primary case. We will use the term imported to specify that the primary case has been infected outside the workplace. A typical scenario is an imported primary case that comes to work (Fig. 2A, red individual). During workdays the primary case will get in proximity contact with other workplace members. These other individuals could get infected and therefore they are potential cases. The number of potential cases will depend on who is the primary case and how many contacts he/she makes during the infectious period. Among those potential cases, some will become infected and they will be counted as secondary cases.

**Figure 2.**
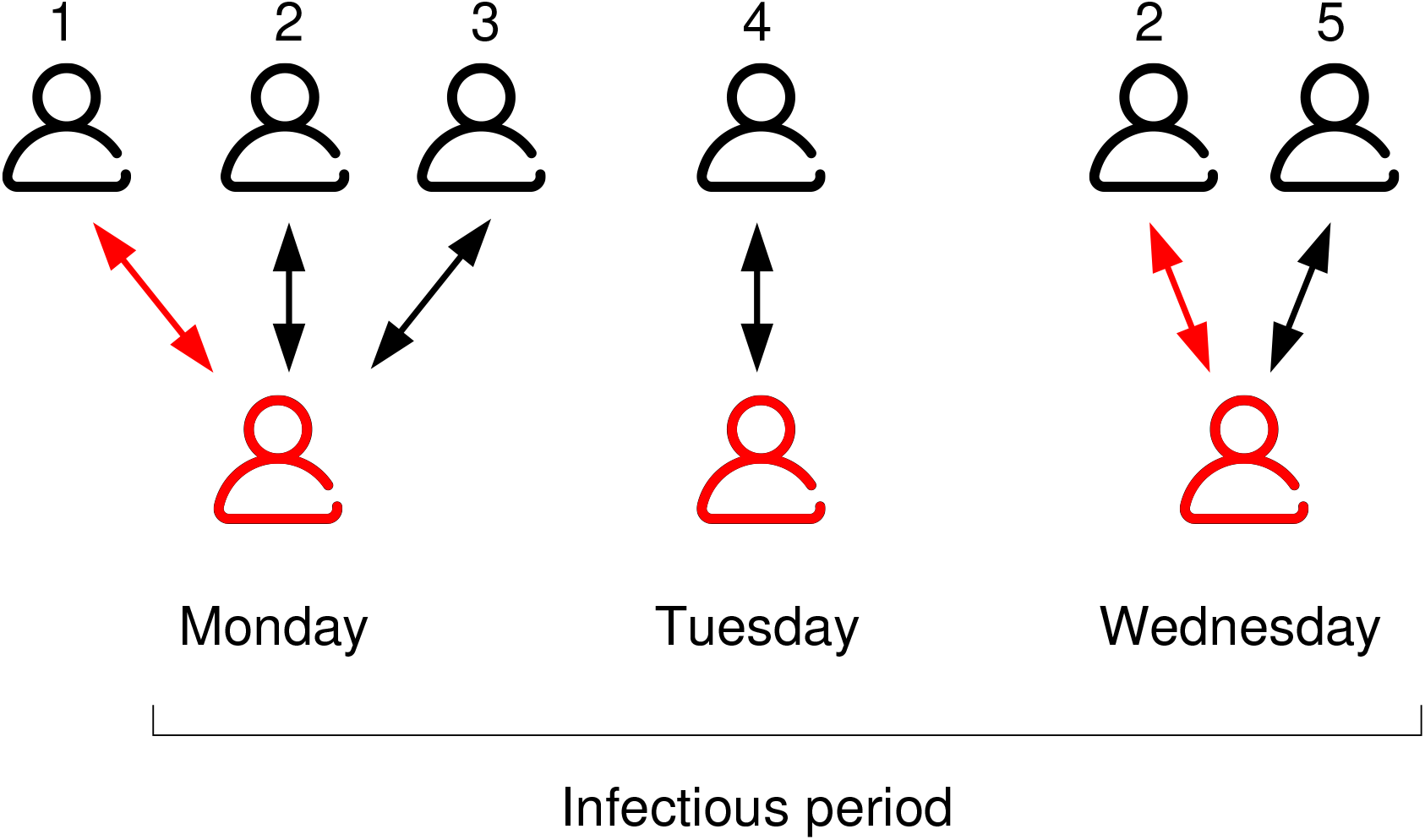
Infection model. A primary imported case (red) comes in contact with other work-place members during the course of the infectious period. Five other individuals are exposed and are potential cases. Disease transmission happens in two instances (red arrows) resulting in two secondary cases.

The more frequent the contacts between a primary case and a potential case are, the higher the chance of disease transmission. We model this aggregate risk of infection using a disease transmission rate per proximity contact *p* and the probability of disease transmission after *c* proximity contacts with an infectious individual

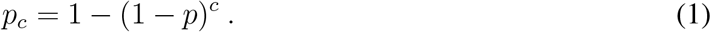

The latter equals 1 minus the probability of no transmission in any of the *c* contacts. In principle we could develop mechanistic models of disease transmission to estimate the transmission rate per contact ^17^. However, that would require us to have a precise knowledge of the environment where these contacts took place (volume, ventilation, temperature), as well as more detailed data of the proximity contacts. Instead, we determine the transmission rate per contact as the value that is consistent with the infectious disease reproductive number.

We developed a procedure to simulate the disease transmission given the proximity contact data, the disease infectious period and the probability of disease transmission after repeated contacts (1). Using this procedure we calculate different statistics characterizing the disease outbreaks within the case-study workplace with the contact records shown in (Fig. 1B). Figure 3 reports the expected reproductive number as a function of the disease transmission rate per contact for infectious periods of 1 or 3 days, which corresponds to the infection periods of the 2019 influenza ^20^ and SARS2 ^21^, respectively. We can invert this relationship to determine the transmission rate per contact that is consistent with a reported reproductive number. Table 1 contains reports of the reproductive number of SARS2, the 2019 influenza and the SARS2 B.1.1.7 variant, together with our estimates of the associated transmission rates. It may sound counterintuitive that the 2019 influenza has a higher estimated transmission rate than SARS2. However, the 2019 influenza generates its reproductive number in a shorter infectious period, which implies a higher transmission rate. More importantly, we estimate a dramatic increase of the transmission rate for the new SARS2 variant, going from 0.014 to 0.041 transmissions per contact.

**Figure 3.**
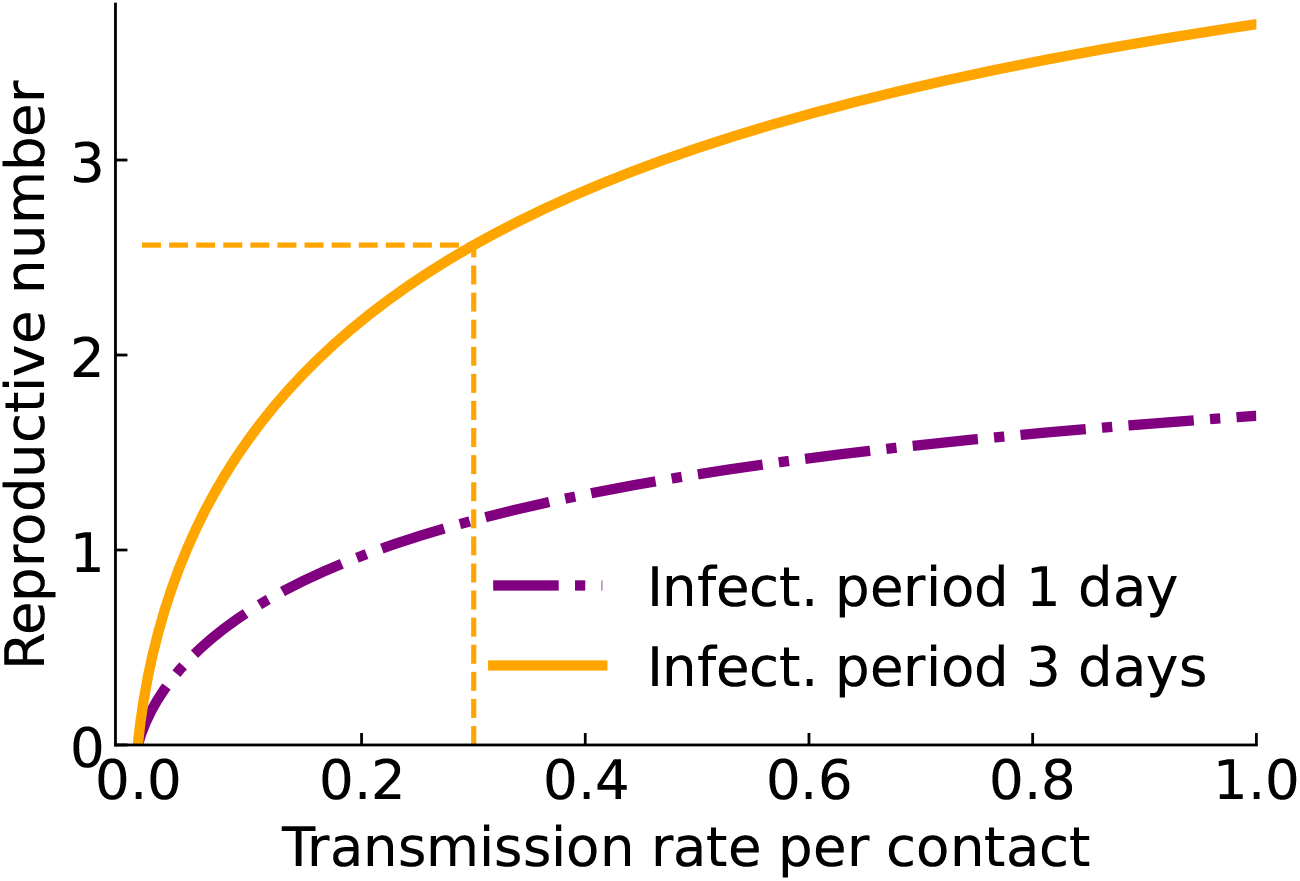
Estimation of the transmission rate per contact: Reproductive number as a function of the transmission rate per proximity contact for an infectious disease with an infectious period of 1 or 3 days. The dashed line connects a given reproductive number with the estimated transmission rate per contact.

**Table 1:** Input and estimated parameters: Infectious disease parameters based on literature reports, together with the estimated transmission rate per contact. Superscripts indicate the reference sources. ^*a*^ Reproductive number corrected for a 20% of contacts taking place at workplace (See Methods). ^*b*^ The reproductive number of SARS2-B.1.1.7 is reported as 1.74 ^8^ or 2.24 ^7^ times higher than of SARS2. We use a factor of 2, which is the average between the two reports. ^*c*^ Assumed equal to the SARS2 value.

| Virus | Reproductive<br>number | Reproductive<br>number, workplace<sup>a</sup> | Infectious<br>period (days) | Transmission<br>rate per contact ($p$) |
| --- | --- | --- | --- | --- |
| 2019 Influenza | 1.7 <sup>19</sup> | 0.34 | 1 <sup>20</sup> | 0.031 |
| SARS-CoV-2 | 2.4 <sup>21</sup> | 0.48 | 3 <sup>21</sup> | 0.014 |
| SARS-CoV-2 New | 4.8 <sup>b</sup> | 1.1 | 3 <sup>c</sup> | 0.041 |

With the estimated disease transmission rate we can run simulations to compare different infectious diseases and to assess the impact of intervention strategies. Mask use is simulated by reducing the transmission rate in proportion to the mask efficiency ^22, 23^. Social distancing is modelled by removing a fraction of the proximity contacts in proportion to the degree of social distancing. The latter yields the same reproductive number as mask use (see Methods) and will not be discussed further.

### Distribution of secondary cases

The distribution of secondary cases contains the information to quantify the rate of super-spreading events. The number of secondary cases is constrained by the number of potential cases, which is determined by the temporal proximity contact network and the infectious period. The distribution of the number of potential cases has an exponential tail (Fig. 4A). The average number of secondary cases generated by an individual is proportional to his/her average number of potential cases (Fig. 4B). This proportionality together with the exponential tail of the distribution of potential cases ultimately results in an exponential tail for the distribution of the number of secondary cases as well (Fig. 4C). The exponential tail is in agreement with reports for the observed distribution of the number of SARS2 secondary infections ^15^. The observed data is best fitted by a negative binomial distribution that is characterized by an exponential tail ^15, 24^.

**Figure 4.**
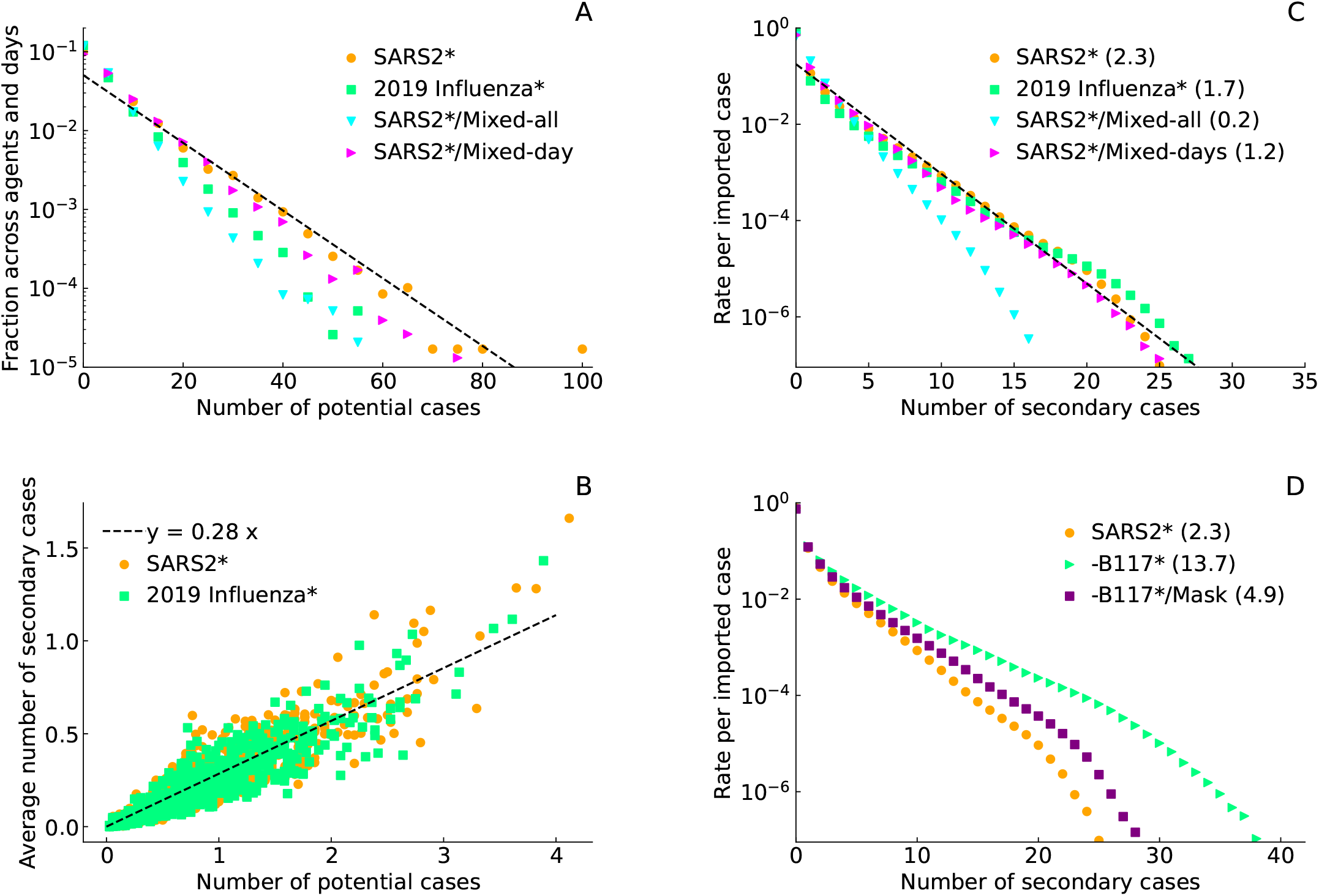
Distribution of potential and secondary cases: A) Distribution of potential cases. B) Average reproductive number of each individual as a function of his/her average number of potential cases. The average is taken over sliding time windows with a size given by the disease infectious period. C) Distribution of secondary cases. The symbol *∗* indicates that these are results from simulations and not real data for the indicated infectious diseases.

Focusing on the number of secondary cases, there is not much difference between the distributions for SARS2 and the 2019 influenza (Fig. 4C). These diseases have a similar propensity to generate super-spreading events. Therefore, the occurrence of super-spreading events is a general property of all infectious diseases transmitted by proximity contacts. The super-spreading events are rooted in the broad distribution of potential cases (Fig. 4A).

Next, we tested the relevance of the temporal structure of the proximity contacts. To this end, we shuffled the data in two different ways. In the most complete randomization, we shuffled the days assigned to all the contacts recorded (mixed-all). In the second approach we keept the temporal cluster within days intact.We permuted days as a block, keeping contacts happening on the same day together (mixed-day). The distributions of the number of potential and of secondary cases have a faster decay for the mixed-all data than for the original records (Fig. 4A,C, circles vs triangles-down). In contrast, the mixed-day shuffling leads to a slight shift from the original distributions (Fig. 4A,C, circles vs triangles-left). The temporal clustering of proximity contacts within days is therefore crucial to obtain the correct distribution of the number of secondary cases. This observation highlights the importance of working with the actual proximity contact records of the target workplace.

### S-index

From the distribution of secondary cases we determine the statistics of super-spreading events. Super-spreading events are defined in different ways in the literature, with different publications having different criteria for how many secondary cases are required for such an event ^24–28^. Here, we define a super-spreading event as an event where the number of secondary cases equals or exceeds 10. Therefore the super-spreading events are associated with the tail of the distribution of the number of secondary cases. The weight of the tail quantifies the rate of super-spreading events per imported case. We name this quantity the *super-spreading index* (*S*-index). The *S*-index will be reported in units of super-spreading events per 1000 imported infectious-disease cases. The *S*-index is determined by both the infection disease and the temporal network of proximity contacts. For example, a pub may have a higher *S*-index than a workplace for the same infectious disease, because proximity contacts are more frequent in a pub than in a workplace.

The legend in Fig. 4C reports the *S*-index for each distribution. The workplace from where the proximity contact data originated has a *S*-index of 2.3 per 1000 imported SARS2 cases. The 2019 influenza has an estimated *S*-index of 1.7, slightly lower than the 2.3 value for SARS2. We also noticed that the full randomization of the day labels (mixed-all) reduced the *S*-index to 0.2 super-spreading events per 1000 imported SARS2 cases. In contrast, the randomization of days keeping records within the same day together (mixed-day) lead to a smaller decrease of the *S*-index from 2.3 to 1.2 super-spreading events per 1000 imported SARS2 cases. Once again, the latter highlights the importance of working with the actual proximity contact records to get the correct estimate of super-spreading rate.

### SARS2-B.1.1.7 variant

The SARS2 B.1.1.7, first detected in the UK, is raising concerns because of its relatively higher reproductive number ^7, 8^. Based on our estimate using the transmission model described above, the transmission rate of the SARS2-B.1.1.7 variant is 0.041 per proximity contact, 3 times larger than the previous SARS2 strain (Table 1). As the transmission rate increases, more potential cases are at risk of becoming actual secondary cases.Therefore, the higher transmission rate results in a wider distribution of the number of secondary cases (Fig. 4D, triangles vs circles). The estimated *S*-index goes up to 13.7 super-spreading events per 1000 imported SARS2-B.1.1.7 cases for the case-study workplace, 6 times higher than the *S*-index for the previous variant. The introduction of mask use, with a simulated mask efficiency of 50% ^29^, brings the estimated *S*-index to 4.9 per 1000 imported SARS2-B.1.1.7 cases, still 2 times larger than the *S*-index for SARS2.

## Discussion

This work highlights the importance of each workplace having an understanding of the proximity contact patterns among its members. The data collection can be conducted without linking the devices’ identification numbers to the individuals’ names to assure anonymity. Given the proximity contact information, each workplace can build its tailored generative model to simulate the spread of SARS2 and the impact of containment strategies. In our opinion this is a requirement to manage infectious disease outbreaks using a distributed approach.

We have estimated the SARS2 transmission rate as 0.014 per contact. We cannot determine if this estimate is specific for the case-study workplace or whether it applies to other workplaces as well. We will be able to answer this question once data from other workplaces is obtained. We also estimated the super-spreading rate in the workplace to be as low as 2.3 super-spreading events per 1000 imported SARS2 cases. The numbers for the SARS2-B.1.1.7 variant are more alarming if the reports regarding the increase in reproductive number prove to be true. The estimated SARS2-B.1.1.7 transmission rate goes up to 0.041 per proximity contact. The super-spreading rate goes up to 13.7 super-spreading events per 1000 imported SARS2-B.1.1.7 cases. Simulated mask use, with 50% mask efficiency, brings the *S*-index down to 4.9 per 1000 imported SARS2-B.1.1.7 cases, still 2 times larger than what is estimated for SARS2.

Our simulations indicate that the 2009 influenza has similar infectious parameters to SARS2. The difference in the reproductive number of SARS2 and the 2009 influenza is rooted in the infectious period, 3 days for SARS2 and 1 day for the 2009 influenza. The applicability of the proposed infection model and the *S*-index extends beyond the ongoing SARS2 outbreak. Our methodology can be applied to simulate the spread of other airborne infectious diseases. The necessary requirements are anonymous proximity contact records and estimates of the disease infectious period and the basic reproductive number in the absence of interventions.

## Methods

### Proximity contact data

The proximity contact data were collected with the social distance monitoring application from the companies Secufy GmbH and Safefactory GmbH. Battery-powered sensors by Secufy are attached to the clothing of individuals and act autonomously without storing data locally. Once activated, each sensor uses Bluetooth Low Energy to measure the signal strength from other sensors. The signal strength is then used to estimate the relative distance to other sensors in its vicinity. If a distance threshold of 1.5 meters is exceeded for a time interval of 15 seconds, the involved sensors beep, directly alerting the affected users that the distance is too close. In addition to this, any shortfall of the minimum distance that lasts longer than 15 seconds is sent to a backend solution from the company Safefactory GmbH, which logs all proximity alarms. The data was collected in a data protection compliant manner at the company ISA S.p.A..

### Infectious disease transmission model

The infectious disease transmission model takes as input the proximity contact records for a workplace with *n* individuals over a period of *m* days. Let *C*_*ijd*_ be the number of contacts between a pair of individuals (*i, j*) on day *d*, where *i* ∈ [1, *…, n*] and *d* ∈ [1, *…, m*]. Following our main model assumption (1), the probability of disease transmission from *i* to *j*, given that *i* becomes infectious on day *d* and remains infectious till *d* + *T*, is given by

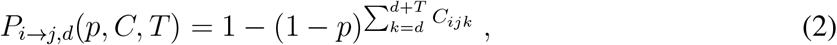

where *i, j* ∈ [1, *…, n*] and *d* ∈ [1, *…, m* − *T*]. The expected reproductive number is given by

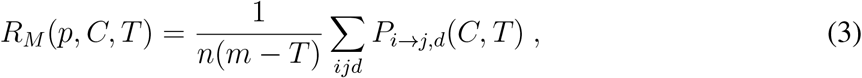

and the distribution of the number of secondary cases (*k*) by

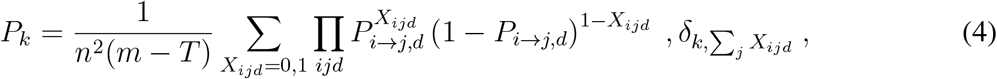

where *δ*_*i,j*_ is the Kronecker delta, *δ*_*i,j*_ = 1 for *i* = *j* and *δ*_*i,j*_ = 0 otherwise. Numerical simulations were performed to generate 10,000 random sets of *X* and calculate *P*_*k*_.

The super-spreading index (*S*-index) is defined as

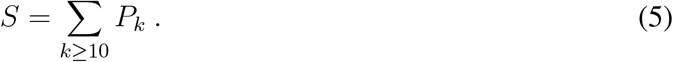

Wearing masks is modelled as a reduction of the probability of disease transmission from p to pµ, where 0 ≤ *µ* ≤ 1 quantifies the mask efficiency.

Social distancing can be modelled by replacing 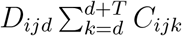 with the binomial sampling

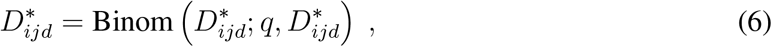

where *q* is the probability that the recorded proximity contacts will take place after social distancing. The expected reproductive number in this case is equal to

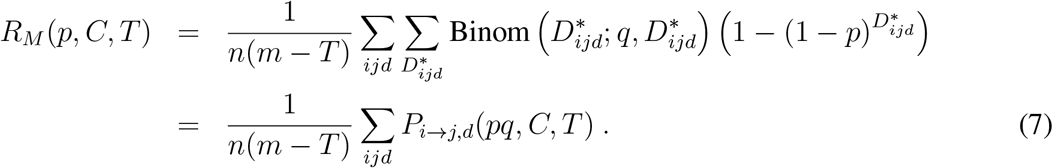

Therefore, social distancing is equivalent to a reduction in the transmission rate from p to pq, which is mathematically equivalent to the mask use model above.

### Estimation of the transmission rate per contact

The transmission rate per contact was estimated as the solution of the equation *R*_*M*_ (*p, C, T*) = *R*_0_, given *C, T* and *R*_0_.

### Reproductive number at the workplace

The percentage of contacts at the work place is reported as 21% ^30^, 25% ^31^, 16% ^32^, and 20%^33^. We assume a value of 20%.

The basic reproductive number of SARS2 in the absence of interventions is reported as 2.4 ^21^. The workplace basic reproductive number therefore equals *R*_0_ = 0.2 × 2.4 = 0.48. The infectious period of SARS2 is 3 days ^21^.

The basic reproductive number of the 2009 influenza in the absence of interventions is reported as 1.7 ^19^. The workplace basic reproductive number therefore equals *R*_0_ = 0.2×1.7 = 0.34. The infectious period of influenza is 1 day ^20^.

The basic reproductive number of the SARS2-B.1.1.7 variant is reported to be 1.74 ^8^ or 2.24 ^7^ higher than for SARS2. We use a factor of 2, which is the average between the two reports, resulting in the reproductive number *R*_0_ = 2.24 × 2.4 = 5.4 and the workplace basic reproductive number *R*_0_ = 0.2 × 2.24 × 2.4 = 1.1. The infectious period of SARS2-B.1.1.7 is assumed to be the same as for SARS2, 3 days.

## Data Availability

Data access is restricted due to intellectual property constraints.

## Acknowledgements

This project was supported by DLR project ResTAT. We thank Secufy GmbH, safefactory GmbH and ISA S.p.A. for providing the proximity contact data and additional information about the study design.

## Author information

DL and EB conceived the project. AV developed the infection transmission model with recommendations from all authors. AV and MS performed data analysis. All authors worked on the manuscript.

## Ethics declaration

ES has indirect shares and convertible loans in Secufy GmbH.

The data we received from ISA contained anonymous identification numbers, one for each individual wearing the buttons. DLR does not have a key to trace back to persons, therefore data for DLR is anonymous. We therefore only process anonymized data.

## Data availability

Data access is restricted due to intellectual property constraints.

## Code availability

Code availability is restricted due to intellectual property constraints.

